# Real-World Effectiveness and Tolerability of Monoclonal Antibodies for Ambulatory Patients with Early COVID-19

**DOI:** 10.1101/2021.03.15.21253646

**Authors:** Brandon J. Webb, Whitney Buckel, Todd Vento, Allison M. Butler, Nancy Grisel, Samuel M. Brown, Ithan D. Peltan, Emily S. Spivak, Mark Shah, Theadora Sakata, Anthony Wallin, Eddie Stenehjem, Greg Poulsen, Joseph Bledsoe

## Abstract

**Importance:** Interventions to reduce hospitalization of patients with COVID-19 are urgently needed. Randomized trials for efficacy suggest that anti-SARS-CoV2 neutralizing monoclonal antibodies (MAb) may reduce medically-attended visits and hospitalization but effectiveness has not been confirmed in a real-world setting.

**Objective:** Estimate the effectiveness of MAb infusion in a real-world cohort of ambulatory patients with early symptomatic COVID-19 at high risk for hospitalization.

**Design:** Quasi-experimental observational cohort study using target trial emulation and causal inference methodology in pre-and post-implementation groups.

**Setting:** Infusion centers and urgent care clinics within an integrated healthcare system in the United States

**Participants:** 13,534 high-risk adult outpatients with symptomatic, laboratory-confirmed COVID-19 within 7 days of symptom onset.

**Exposures:** A single intravenous infusion of either bamlanivimab 700 mg or casirivimab/imdevimab 1200 mg/1200 mg.

**Main Outcomes and Measures:** The primary outcome was emergency department visit or hospitalization within 14 days of positive test. Patients who received MAb infusion were compared to contemporaneous controls using inverse probability of treatment weighting, and to a pre-implementation cohort using propensity-weighted interrupted time series analysis. An exploratory analysis compared effectiveness of casirivimab/imdevimab and bamlanivimab.

**Results:** 7404 patients who would have been MAb-eligible were identified in a pre-implementation cohort (July 1-November 27, 2020). In the post-implementation period (November 28, 2020-January 28, 2021), 594 received MAb treatment and 5536 MAb-eligible patients did not. Among Mab recipients, 479 (80.6%) received bamlanivimab and 115 (19.4%) casirivimab/imdevimab. The primary outcome occurred in 75 (12.6%) MAb recipients, 1018 (18.4%) contemporaneous controls, and 1525 (20.6%) patients in the pre-implementation cohort. MAb treatment was associated with fewer subsequent emergency department visits and hospitalizations (odds ratio estimating the average treatment effect 0.69, 95% CI 0.60-0.79). After implementation, propensity-weighted probability of emergency department visit or hospitalization decreased by 0.7% per day (95% CI 0.03-0.10%, p<0.001). Overall, 7 (1.2%) MAb patients experienced an adverse event; two (0.3%) were considered serious. In the exploratory analysis, the effect of casirivimab/imdevimab versus bamlanivimab was not significant (OR 0.52, 95% CI 0.17-1.63, p=0.26).

**Conclusions and Relevance:** MAb treatment of high-risk ambulatory patients with early COVID-19 was well-tolerated and effective at preventing the need for subsequent medically-attended care.

**Key Points Section:** *Question:* What is the real-world effectiveness of COVID-19 monoclonal neutralizing antibody (MAb) infusions in high-risk, ambulatory patients?

*Findings:* 594 high-risk, early-symptomatic adults with COVID-19 treated with MAb infusion were compared to 5536 contemporaneous controls using inverse probability of treatment weighting, and to 7404 patients in a pre-implementation cohort using propensity-weighted interrupted time series analysis. MAb treatment was associated with fewer subsequent emergency department visits and hospitalizations (odds ratio 0.69 (95% CI 0.60-0.79). After MAb implementation the probability of emergency department visit or hospitalization decreased by 0.7% per day, 95% CI 0.03-0.10%, p<0.001).

*Meaning:* Monoclonal antibody infusion within seven days of symptom onset in high-risk ambulatory adults with COVID-19 appears to prevent subsequent emergency department visits and hospitalization. Further evaluation of the differences between specific Mab products is warranted.

## Introduction

Monoclonal antibodies (MAbs) designed to avidly bind to the receptor binding domain of the SARS-CoV-2 spike glycoprotein are an emerging passive neutralizing therapy for COVID-19. In fall 2020, bamlanivimab (Eli Lilly, Indianapolis IN) and casirivimab/imdevimab (Regeneron, Tarrytown NY) received United States Food and Drug Administration emergency use authorization (EUA) for treatment of mild/moderate symptomatic COVID-19 in ambulatory patients at higher risk for hospitalization^1,2^ and were distributed for administration under a Department of Health and Human Services program. Secondary endpoints from phase II/III clinical trials suggest that early administration of these MAbs may prevent emergency department (ED) visits or hospitalization, particularly in highest-risk subgroups.^3-5^ However, additional evidence is needed to clarify the true magnitude of this effect and better characterize adverse event rates. Here, we apply principles of target trial emulation and causal-inference methodology to assess the real-world effectiveness and tolerability of MAbs for COVID-19 after implementation in a large, integrated healthcare system.

## Methods

### Setting and Data Repository

Intermountain Healthcare is a regional integrated healthcare system that provides care to more than 1.5 million patients each year. During the COVID-19 pandemic, Intermountain has offered SARS-CoV-2 testing at urgent care facilities, emergency departments, and community drive-up testing sites in Utah and southeastern Idaho.As part of patient notification and clinical trial enrollment processes, a report of all positive test results is generated daily and includes clinical and demographic data. Clinical for this study were extracted from the Intermountain enterprise data warehouse and Intermountain Prospective Observational COVID-19 (IPOC) database. Comorbidities were defined using the Charlson and Elixhauser definitions.^6,7^ Comorbidity data were complete for patients with prior encounters within the integrated health system, patients with missing prior encounter data were excluded.

### Monoclonal Antibody Eligibility and Delivery

At the time of EUA approval, due to high community transmission, patients eligible for MAb using the EUA criteria far exceeded infusion capacity. To address resource scarcity, the Scarce Medications Allocation Subcommittee of the Utah Crisis Standards of Care (CSC) Workgroup was convened with the dual aims of targeting available MAb infusions to patients most likely to benefit and ensuring equity in delivery.^8^ A simple clinical prediction score for predicting severe COVID-19 resulting in hospitalization or mortality among ambulatory patients was validated in a large cohort of Utah patients and adopted for use in MAb allocation.^9^ The score weights age, gender, shortness of breath, comorbidities, and non-white race or Hispanic/Latinx ethnicity to address recognized disparities in poor COVID-19 outcomes in these populations.

Clinical eligibility for MAb was defined by the CSC committee as the following: 1) at or above the risk score threshold (set at ≥7.5 points, which identified approximately the top decile of estimated risk among COVID-19 positive patients), 2) laboratory-confirmed COVID-19 by nucleic acid amplification test or antigen detection, 3) symptomatic disease with symptom onset within no more than 7 days. Patients were ineligible to receive MAb therapy for the following conditions: 1) hospitalized due to COVID-19, 2) new, COVID-related hypoxemia (defined as peripheral oxygen saturation <90% at rest or new supplemental oxygen requirement, or for those with chronic hypoxia, a new change in baseline saturation or oxygen demand, 3) pregnant, or 4) known hypersensitivity to other monoclonal drugs. Eligibility criteria for pediatric patients differed from adults and patients <18 years of age are not included in this study.

To provide equitable access to treatment, 16 regional infusion sites (7 hospital-based infusion centers and 9 urgent care facilities) were selected based on population density, prevalence of underserved patients, and travel time for patients in rural communities. Two pathways for patient identification were developed: first, a hotline and email address were set up to receive referrals from patients and providers in the state. Second, recognizing that patients with poorer healthcare access might have less awareness about treatment availability and less likely to have a referring provider, a process was implemented to proactively identify patients at the time of test positivity. To do this, the risk calculator result was electronically integrated into the daily report of all new positive cases. Each day, a clinician using the report reviewed available medical records for candidate patients with new positive tests and attempted to contact patients by telephone to verify eligibility for MAb infusion. A telephone interpreter service was used for all patients with non-English language preferences. Eligible patients were then scheduled at the nearest infusion site with appointment availability.

At a daily huddle, all infusion sites reported infusion-associated adverse events, defined as symptoms requiring clinical evaluation or management during the infusion or one-hour observation period. The first bamlanivimab infusions were provided in infusion centers and urgent care clinics on December 1, 2020 (to patients with positive tests as of November 28, 2020) and the first casirivimab/imdevimab infusions were administered on December 30, 2020. Due to drug preparation requirements, when casirivimab/imdevimab became available it replaced bamlanivimab as the sole product used at infusion centers, whereas urgent care sites continued to administer only bamlanivimab.

### Study Design and Statistical Analysis

To minimize bias and improve causal inference in an observational study design, we applied principles of target trial emulation in designing our analyses.^10^ First, we identified two cohorts: 1) a pre-implementation cohort comprised of ambulatory patients with positive COVID-19 tests performed between July 1 and November 27, 2020 and 2) a post-implementation cohort of patients with positive tests during the period when MAb therapy was available (November 28, 2020 through January 28, 2021). All patients had at least 14 days of follow-up from the time of testing. We limited the entire cohort to patients who would have been screened for MAb eligibility based on a risk prediction score of at least 7.5 points, calculated using the same electronic method applied in the actual patient identification process. We then excluded patients who were either admitted at the time of COVID-19 testing or within 72 hours following testing. By doing this, we excluded patients who would not have been eligible for MAbs because of hospitalization, or advanced disease and hypoxia at the time of screening, and those who would not have had sufficient time to receive MAb treatment and derive benefit had treatment been available.

We prespecified analyses in the pre- and post-implementation cohorts using causal inference methodology intended to address different biases in observational data in a complementary way. The primary outcome was a composite of subsequent ED visit or hospitalization in the 14 days after testing. This outcome was chosen to account for increasing admission thresholds during the study period, which presented risk of secular confounding affecting the pre-implementation cohort. Admission criteria were static during the period in which MAbs were available, and we performed a sensitivity analysis with the outcome of hospitalization within 14 days in the post-implementation group.

To estimate the average treatment effect in the treated (ATT) for patients who received MAbs, we first developed a propensity model using multivariable logistic regression to estimate the propensity for MAb treatment within the post-implementation group.^11^ Propensity model variables were selected on the basis of expert opinion, previous evidence, plausibility, and results of a focus group of clinicians involved in the MAb screening process. We then used the propensity model to conduct inverse probability of treatment weighting (IPTW) to estimate the ATT for the primary outcome associated with MAb treatment.^12^ Adequate covariate balance after IPTW was assessed using standardized mean differences. We performed a sensitivity analysis limited to the 14-day hospitalization outcome only. Adjustment for multiple comparisons was not performed for the sensitivity analysis and results should be considered hypothesis generating. To aid in comparing these results to those from randomized trials, we used the Chatellier method of estimating the number needed to treat (NNT) from the odds ratio.^13^ We then conducted an interrupted time series (ITS) analysis using segmented regression with propensity-weighting to estimate the change in probability of the primary outcome in patients before and after MAb treatment was available.^14^ The segmented regression for ITS analysis was conducted at the individual patient level with days as the time series unit, applying IPTW using the propensity model developed in the post-implementation cohort. The output of the model includes the baseline trend in the pre-implementation group, and the level change and trend change post-implementation in the per-day probability of subsequent emergency department visit or hospitalization.

Finally, we conducted a planned secondary analysis to compare the efficacy of casirivimab/imdevimab with bamlanivimab. We took advantage of the natural experiment resulting from complete transition of infusion centers from bamlanivimab to casirivimab/imdevimab therapy once the latter agent became available. Analyses were restricted to patients who received their treatment at infusion centers and adjusted for residual unbalanced features and secular trend in non-treated contemporaneous controls in a multivariable regression model. Statistical analyses were performed using R, version 4.0.3 (Vienna, Austria). Study elements fulfilled STROBE reporting guidelines for cohort studies. This study was determined to be exempt from review by the Intermountain Institutional Review Board.

## Results

A total of 13,534 ambulatory patients with symptomatic, laboratory-confirmed COVID-19 with a risk score ≥7.5 points were included in the study, including 7,404 patients in the pre-implementation period and 6,130 in the post-implementation period (See Figure 1). Of the latter, 594 (9.7%) of patients received a monoclonal antibody infusion; 479 (80.6%) received bamlanivimab and 115 (19.4%) received casirivimab/imdevimab. Demographic and clinical characteristics by group are displayed in Table 1. Urgent care was the most common infusion site (n=317 [53%]), followed by infusion centers (272 [46%]) and one rural emergency department (5 [1%]). Median time from SARS-CoV-2 test sample collection to infusion was 53 hours (IQI 49-74). Among patients who received MAb treatment, the mean age was 65 years (SD 13) and median number of comorbidities was 5 (interquartile interval (IQI) 3-6), of which obesity 397 (67%), diabetes mellitus 390 (66%) and chronic pulmonary disease 347 (58%) were common. Factors included in the propensity model are listed in Supplementary eTable 1. IPTW effectively reduced imbalance in the treated versus contemporaneous controls as demonstrated by the standardized mean differences plot (Supplementary eFigure 1).

**Table 1.**
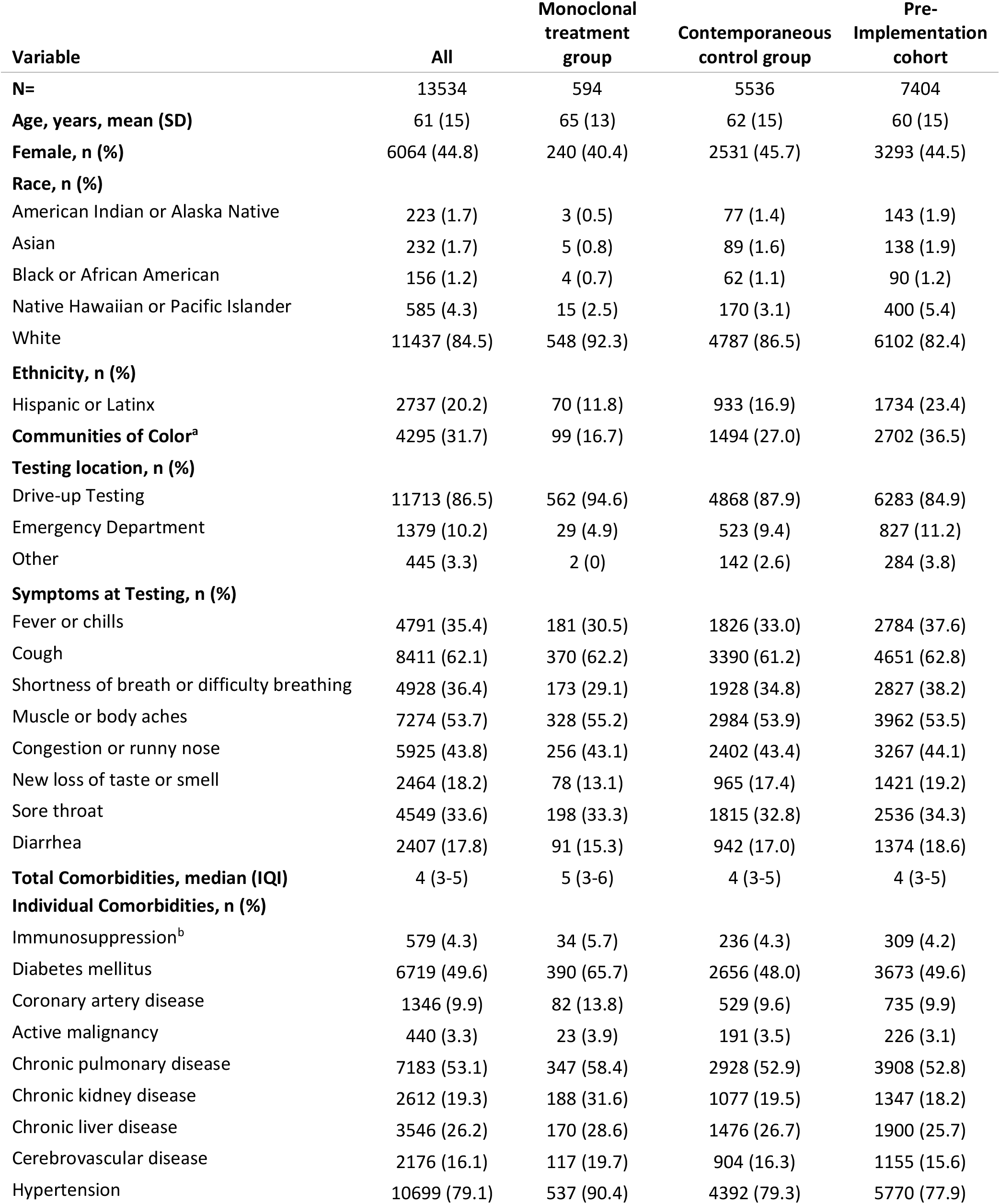
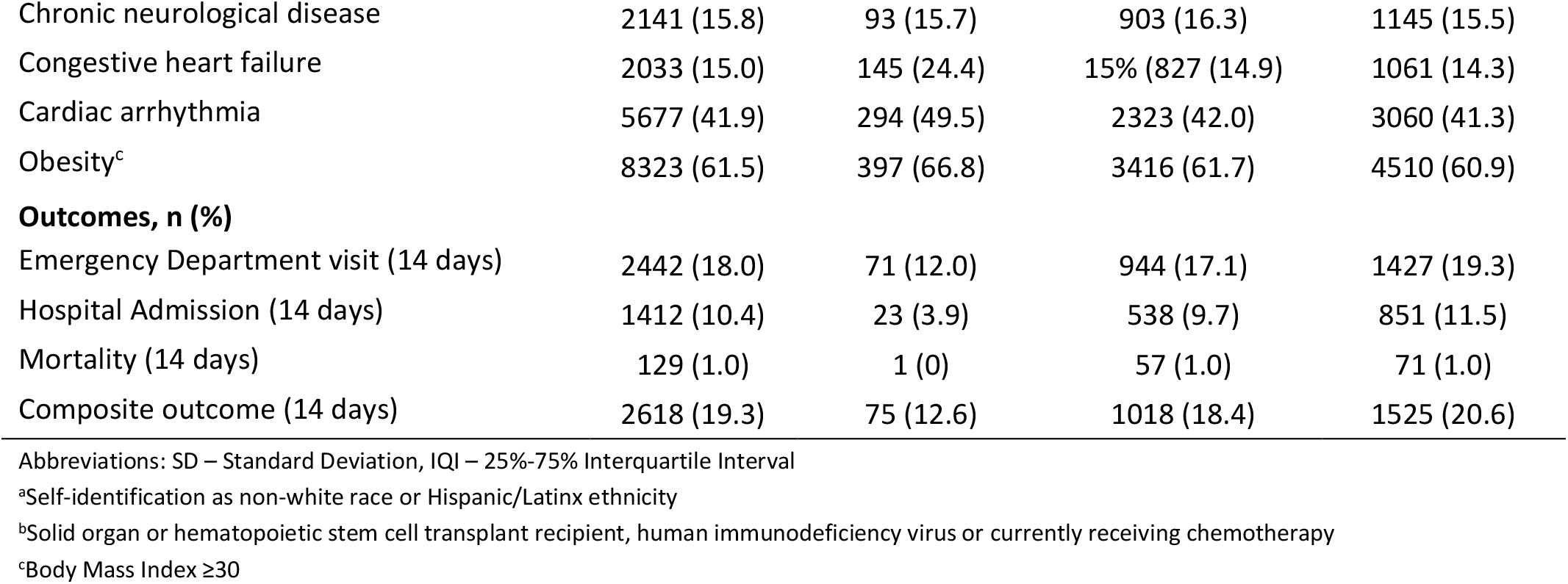
**Clinical Features by Treatment and Non-Treatment Groups**

| Variable | All | Monoclonal<br>treatment<br>group | Contemporaneous<br>control group | Pre-Implementation<br>cohort |
| --- | --- | --- | --- | --- |
| **N=** | 13534 | 594 | 5536 | 7404 |
| **Age, years, mean (SD)** | 61 (15) | 65 (13) | 62 (15) | 60 (15) |
| **Female, n (%)** | 6064 (44.8) | 240 (40.4) | 2531 (45.7) | 3293 (44.5) |
| **Race, n (%)** |  |  |  |  |
| American Indian or Alaska Native | 223 (1.7) | 3 (0.5) | 77 (1.4) | 143 (1.9) |
| Asian | 232 (1.7) | 5 (0.8) | 89 (1.6) | 138 (1.9) |
| Black or African American | 156 (1.2) | 4 (0.7) | 62 (1.1) | 90 (1.2) |
| Native Hawaiian or Pacific Islander | 585 (4.3) | 15 (2.5) | 170 (3.1) | 400 (5.4) |
| White | 11437 (84.5) | 548 (92.3) | 4787 (86.5) | 6102 (82.4) |
| **Ethnicity, n (%)** |  |  |  |  |
| Hispanic or Latinx | 2737 (20.2) | 70 (11.8) | 933 (16.9) | 1734 (23.4) |
| **Communities of Color<sup>a</sup>** | 4295 (31.7) | 99 (16.7) | 1494 (27.0) | 2702 (36.5) |
| **Testing location, n (%)** |  |  |  |  |
| Drive-up Testing | 11713 (86.5) | 562 (94.6) | 4868 (87.9) | 6283 (84.9) |
| Emergency Department | 1379 (10.2) | 29 (4.9) | 523 (9.4) | 827 (11.2) |
| Other | 445 (3.3) | 2 (0) | 142 (2.6) | 284 (3.8) |
| **Symptoms at Testing, n (%)** |  |  |  |  |
| Fever or chills | 4791 (35.4) | 181 (30.5) | 1826 (33.0) | 2784 (37.6) |
| Cough | 8411 (62.1) | 370 (62.2) | 3390 (61.2) | 4651 (62.8) |
| Shortness of breath or difficulty breathing | 4928 (36.4) | 173 (29.1) | 1928 (34.8) | 2827 (38.2) |
| Muscle or body aches | 7274 (53.7) | 328 (55.2) | 2984 (53.9) | 3962 (53.5) |
| Congestion or runny nose | 5925 (43.8) | 256 (43.1) | 2402 (43.4) | 3267 (44.1) |
| New loss of taste or smell | 2464 (18.2) | 78 (13.1) | 965 (17.4) | 1421 (19.2) |
| Sore throat | 4549 (33.6) | 198 (33.3) | 1815 (32.8) | 2536 (34.3) |
| Diarrhea | 2407 (17.8) | 91 (15.3) | 942 (17.0) | 1374 (18.6) |
| **Total Comorbidities, median (IQR)** | 4 (3-5) | 5 (3-6) | 4 (3-5) | 4 (3-5) |
| **Individual Comorbidities, n (%)** |  |  |  |  |
| Immunosuppression<sup>b</sup> | 579 (4.3) | 34 (5.7) | 236 (4.3) | 309 (4.2) |
| Diabetes mellitus | 6719 (49.6) | 390 (65.7) | 2656 (48.0) | 3673 (49.6) |
| Coronary artery disease | 1346 (9.9) | 82 (13.8) | 529 (9.6) | 735 (9.9) |
| Active malignancy | 440 (3.3) | 23 (3.9) | 191 (3.5) | 226 (3.1) |
| Chronic pulmonary disease | 7183 (53.1) | 347 (58.4) | 2928 (52.9) | 3908 (52.8) |
| Chronic kidney disease | 2612 (19.3) | 188 (31.6) | 1077 (19.5) | 1347 (18.2) |
| Chronic liver disease | 3546 (26.2) | 170 (28.6) | 1476 (26.7) | 1900 (25.7) |
| Cerebrovascular disease | 2176 (16.1) | 117 (19.7) | 904 (16.3) | 1155 (15.6) |
| Hypertension | 10699 (79.1) | 537 (90.4) | 4392 (79.3) | 5770 (77.9) |
| Chronic neurological disease | 2141 (15.8) | 93 (15.7) | 903 (16.3) | 1145 (15.5) |
| Congestive heart failure | 2033 (15.0) | 145 (24.4) | 15% (827 (14.9) | 1061 (14.3) |
| Cardiac arrhythmia | 5677 (41.9) | 294 (49.5) | 2323 (42.0) | 3060 (41.3) |
| Obesity<sup>c</sup> | 8323 (61.5) | 397 (66.8) | 3416 (61.7) | 4510 (60.9) |
| **Outcomes, n (%)** |  |  |  |  |
| Emergency Department visit (14 days) | 2442 (18.0) | 71 (12.0) | 944 (17.1) | 1427 (19.3) |
| Hospital Admission (14 days) | 1412 (10.4) | 23 (3.9) | 538 (9.7) | 851 (11.5) |
| Mortality (14 days) | 129 (1.0) | 1 (0) | 57 (1.0) | 71 (1.0) |
| Composite outcome (14 days) | 2618 (19.3) | 75 (12.6) | 1018 (18.4) | 1525 (20.6) |

Abbreviations: SD – Standard Deviation, IQI – 25%-75% Interquartile Interval
<sup>a</sup>Self-identification as non-white race or Hispanic/Latinx ethnicity
<sup>b</sup>Solid organ or hematopoietic stem cell transplant recipient, human immunodeficiency virus or currently receiving chemotherapy
<sup>c</sup>Body Mass Index $\geq 30$

**Figure 1.**
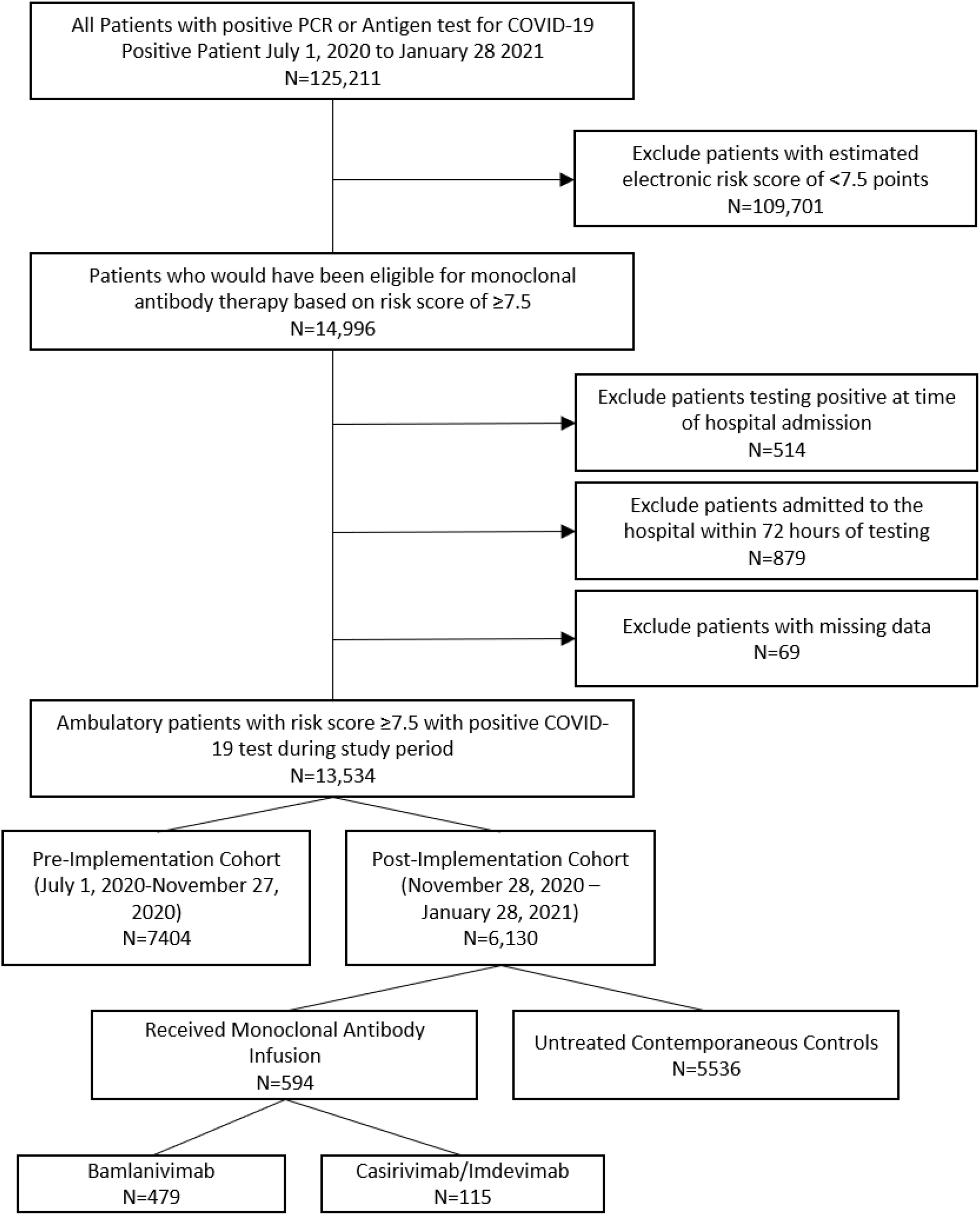
CONSORT-style flow diagram.

The primary outcome occurred in 75 (12.6%) patients in the MAb group, 1018 (18.4%) in the contemporaneous controls and 1525 (20.6%) in the pre-implementation cohort (Table 1). Subsequent ED visits occurred in 71 (12%) patients in the MAb treatment group compared to 944 (17%) in the contemporaneous non-treated group and1427 (19%) in the pre-implementation cohort. In the MAb treatment group, 23 (3.9%) patients were admitted to the hospital within 14 days of testing compared to 538 (9.7%) in the contemporaneous controls and 851 (11.5%) in the pre-implementation cohort.

After IPTW, MAb-treated patients were significantly less likely to have an ED visit or hospitalization within 14 days of testing compared to contemporaneous controls (odds ratio estimating the ATT 0.69, 95% CI 0.60-0.79, p<0.001). This corresponds to an estimated number NNT to prevent one medically attended visit of 7.6. In the sensitivity analysis, the odds of 14-day hospital admission were also significantly decreased in MAb-treated patients (OR 0.43, 95% CI 0.35-0.53, p<0.001). In the interrupted time series analysis, propensity-weighted probability of ED visit or hospitalization decreased by 0.7% per day (CI 0.003-0.10%, p<0.001) after implementation of MAb treatment (Figure 2, eTable 3).

**Figure 2.**
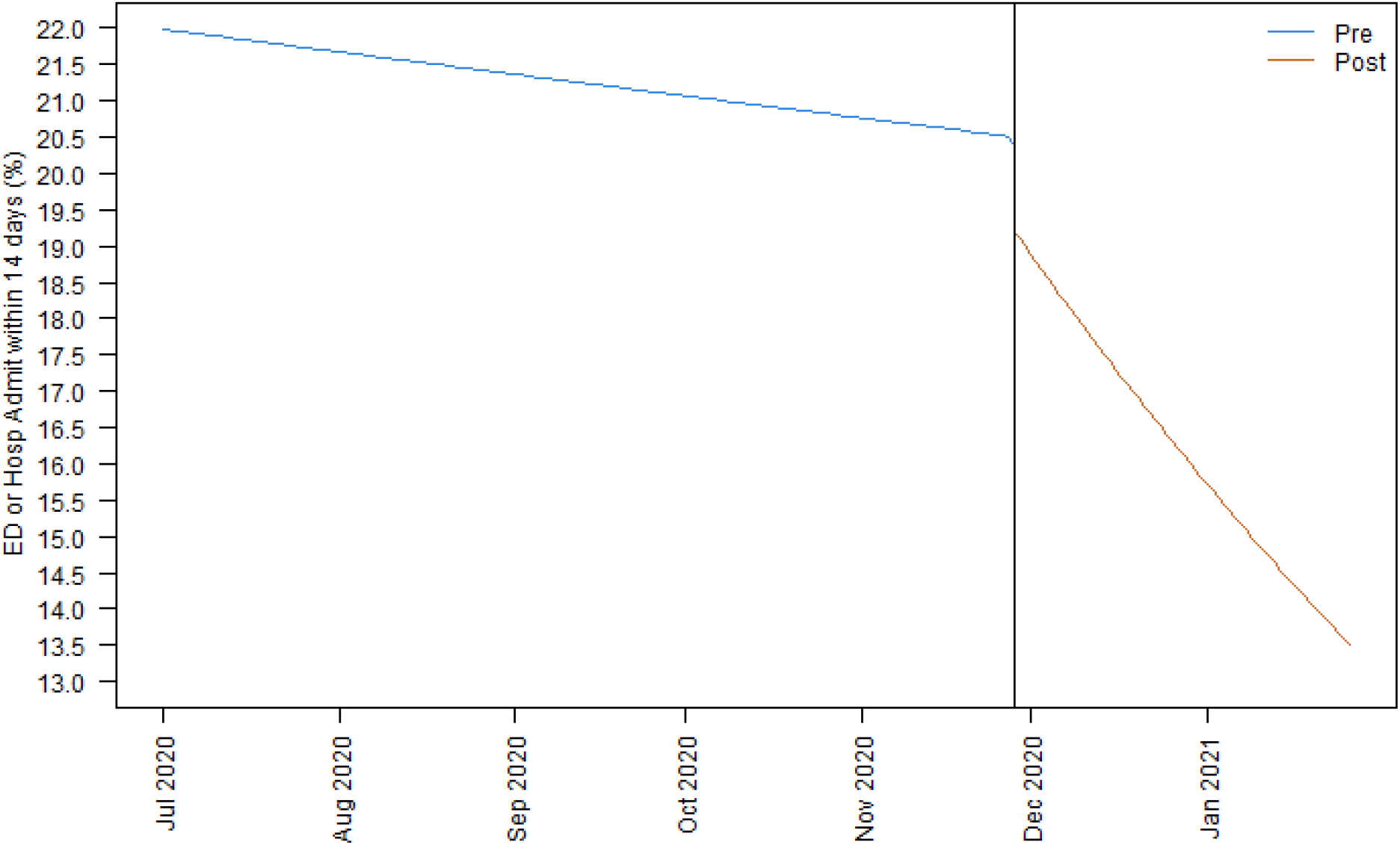
**Interrupted time series analysis estimating differences in the daily probability of emergency department visit or hospital admission pre- and post-implementation of monoclonal therapy**

In a subset of MAb-treated patients who received their treatment at an infusion center, (n=272), 157 (55.5%) received bamlanivimab and 115 (44.5%) received casirivimab/imdevimab (Supplementary eTable 2). Features were generally comparable between groups with the exception of modest differences in obesity and female gender. Of patients receiving casirivimab/imdevimab, 10 (8.7%) met the composite endpoint, compared to 19 (12.1%) patients who received bamlanivimab. In the casirivimab/imdevimab group, 1 (0.9%) was hospitalized within 14 days, compared to 7 (4.5%) patients infused with bamlanivimab (Supplementary Table 1). Using logistic regression adjusting for gender, obesity and the secular trend in non-treated contemporaneous controls (eTable 4), odds ratio for the primary endpoint for casirivimab/imdevimab versus bamlanivimab was 0.52 (95% CI 0.17-1.63, p=0.26).

Both monoclonal antibody products were well tolerated. A total of 7 (1.2%) patients experienced infusion-associated adverse events (Table 3). Two events (0.3%) were considered severe – one patient with known coronary disease developed chest pain during infusion and another had a syncopal episode; both patients were managed in the emergency department with good outcomes. Five (0.8%) mild reactions were observed: pruritis, hives, rigors, nausea/vomiting and oral tingling. None of the mild reactions required premature discontinuation of the infusion and all resolved within hours to days.

**Table 2.** Clinical Features by Monoclonal Product.

| **Variable** | **All** | **Bamlanivimab** | **Casirivimab/Imdevimab** |
| --- | --- | --- | --- |
| N= (%) | 594 | 479 (80.6) | 115 (19.4) |
| **Age, years, mean (SD)** | 65 (13) | 65 (13) | 66 (15) |
| **Female, n (%)** | 240 (40.4) | 186 (38.8%) | 54 (47.0) |
| **Race, n (%)** |  |  |  |
| American Indian or Alaska Native | 3 (0.5) | 3 (0.6) | 0 (0) |
| Asian | 5 (0.8) | 4 (0.8) | 1 (0.9) |
| Black or African American | 4 (0.7) | 3 (0.6) | 1 (0.9) |
| Native Hawaiian or Pacific Islander | 15 (2.5) | 14 (2.9) | 1 (0.9) |
| White | 548 (92.3) | 440 (91.9) | 108 (93.9) |
| **Ethnicity** |  |  |  |
| Hispanic or Latinx | 70 (11.8) | 55 (11.5) | 15 (13.0) |
| **Communities of Color<sup>a</sup>** | 99 (16.7) | 80 (16.7) | 19 (16.5) |
| **Total Comorbidities, median (IQR)** | 5 (3-6) | 5 (3-6) | 4 (3-5) |
| **Individual comorbidities, n (%)** |  |  |  |
| Immunosuppression<sup>b</sup> | 34 (5.7) | 28 (5.8) | 6 (5.2) |
| Diabetes mellitus | 390 (65.7) | 317 (66.2) | 73 (63.5) |
| Coronary artery disease | 82 (13.8) | 66 (13.8) | 16 (13.9) |
| Active malignancy | 23 (3.9) | 17 (3.5) | 6 (5.2) |
| Chronic pulmonary disease | 347 (58.4) | 282 (58.9) | 65 (56.5) |
| Chronic kidney disease | 188 (31.6) | 153 (31.9) | 35 (30.4) |
| Chronic liver disease | 170 (28.6) | 136 (28.4) | 34 (29.6) |
| Cerebrovascular disease | 20% (117 (19.7)) | 97 (20.3) | 20 (17.4) |
| Hypertension | 537 (90.4) | 435 (90.8) | 102 (88.7) |
| Chronic neurological disease | 0 (0) | 0 (0) | 0 (0) |
| Congestive heart failure | 145 (24.4) | 126 (26.3) | 19 (16.5) |
| Cardiac arrhythmia | 294 (49.5) | 237 (49.5) | 57 (49.6) |
| Obesity<sup>c</sup> | 397 (66.8) | 335 (69.9) | 62 (53.9) |
| Hours from test to infusion, median (IQR) | 53 (49-74) | 54 (49-74) | 52 (44-74) |
| **Infusion Location** |  |  |  |
| Emergency department | 5 (0.8) | 5 (10.4) | 0 (0) |
| Infusion Center | 272 (45.8) | 157 (32.8) | 115 (100) |
| Urgent Care | 317 (53.4) | 317 (66.2) | 0 (0) |
| **Infusion-associated Adverse Events** |  |  |  |
| Any | 7 (1.2) | 6 (1.3) | 1 (0.9) |
| Mild | 5 (0.8) | 4 (0.8) | 1 (0.9) |
| Severe<sup>d</sup> | 2 (0.3) | 2 (0.4) | 0 (0) |
| **Outcomes** |  |  |  |
| Emergency department visit (14 days) | 71 (12.0) | 62 (12.9) | 9 (7.8) |
| Hospital admission (14 days) | 23 (3.9) | 22 (4.6) | 1 (0.9) |
| Mortality (14 days) | 1 (0.2) | 1 (0.2) | 0 (0) |
| Composite outcome (14 days) | 75 (12.6) | 65 (13.6) | 10 (8.7) |

Abbreviations: SD – Standard Deviation, IQI – 25%-75% Interquartile Interval
<sup>a</sup>Self-identification as non-white race or Hispanic/Latinx ethnicity
<sup>b</sup>Solid organ or hematopoietic stem cell transplant recipient, human immunodeficiency virus or currently receiving chemotherapy
<sup>c</sup>Body Mass Index $\geq 30$
<sup>d</sup>Severe adverse events defined as requiring referral to emergency department for management

**Table 3.** Infusion-associated adverse events.

| **Reported Symptom** | **Drug** | **Outcome** | **Serious Adverse Event?** |
| --- | --- | --- | --- |
| Chest pain | Bamlanivimab | Known coronary disease, resolved with anti-angina treatment in the emergency department | Yes |
| Oral tingling | Bamlanivimab | Self-limited | No |
| Pruritis without rash | Bamlanivimab | Infusion paused and safely resumed, treated with diphenhydramine | No |
| Hives | Casirivimab/Imdevimab | Completed infusion, treated with diphenhydramine and short course of oral methylprednisolone | No |
| Rigors | Bamlanivimab | Infusion paused, resumed a slower rate, no hypotension | No |
| Nausea and emesis | Bamlanivimab | Completed infusion, palliated with ondansetron, persistent nausea lasted 4 days | No |
| Syncope | Bamlanivimab | Infusion terminated, patient was triaged to the emergency department and diagnosed with likely vasovagal etiology, no anaphylactoid features | Yes |

## Discussion

In this real-world evaluation of SARS-CoV-2 monoclonal antibody infusion therapy, administration of MAb treatment to a high-risk population of ambulatory COVID-19 patients within seven days of symptom onset was associated with significant reductions in subsequent emergency department visits and hospital admissions. For context, the NNT to prevent one medically-attended visit was less than 8. This effect estimate is consistent with sub-group analyses of higher-risk patients from MAb clinical trials,^3-5^ supporting the clinical effectiveness of these drugs and informing optimal patient selection for treatment. It is likely that the estimated effect size in our cohort was due to enrichment for a population both at greater risk for poor outcomes and earlier in the symptom course than specified by the EUA criteria.^1,2^

These results lend additional support to the concept that passive immune therapies^15^ are effective when administered early after symptom onset when viral replication is highest^16^ and in patients who fail to mount a robust early humoral response.^5^ Of note, while the EUA for both agents authorizes administration for up to 10 days after symptom onset,^1,2^ symptom onset beyond day 7 was an exclusion criterion in the casirivimab/imdevimab trial,^5^ and the median time from symptom onset to infusion in the bamlanivimab trial^3^ was 4 days. Operationally, we found that because of test-seeking behaviors, scheduling the monoclonal antibody infusion within two days of testing was necessary to achieve infusion within this early window. We employed centralized eligibility review and recruitment paired with a network of infusion locations to identify MAb-eligible patients and meet symptom-to-infusion interval goals.

Our patients’ experience also corroborates safety data from clinical trials, suggesting that MAb treatment is well-tolerated. Serious events were rare and no anaphylactic-type events were observed. However, data from much larger populations will be necessary to accurately characterize the incidence of rare events. Although we observed a signal suggesting a possible efficacy difference favoring casirivimab/imdevimab over bamlanivimab in reducing medically attended visits, our sample size was inadequate to confirm. This merits further comparative effectiveness investigation in larger cohorts. Early clinical trial data suggest that the efficacy of combination monoclonal antibodies targeting multiple, non-overlapping epitopes may be superior to monovalent products.^4^ Differential activity against emerging variant SARS-CoV2 strains may also play a role. In contrast to casirivimab/imdevimab, bamlanivimab may exhibit reduced activity against the E484^17^ mutation found in the B.1.351, P.1 and B.1.526 strains and perhaps the L452 mutation found in the B.1.149 California variant.^18^ At the time of submission, the Utah Public Health Laboratory had reported sequencing 8951 isolates of which 112 (1.3%) were identified as B.1.149, 67 (0.8%) B.1.1.7 and none as B.1.351 or P.1.^19^

Our study has several limitations. First, despite a target trial emulation design and complementary causal inference analyses, it is impossible to fully mitigate biases inherent to observational data. We recognize that in the post-implementation group, unmeasurable or unmeasured confounding factors may influence the estimates for counterfactual treatment effect. This is addressed by the interrupted time series analysis, and we are reassured that this complementary analysis does corroborate the ATT estimate. However, it is possible that unmeasured factors influenced these estimates. Although such interventions are uncommon for ambulatory patients in our health system, we were unable to measure concomitant prescription of other repurposed therapies such as corticosteroids. In addition, date of symptom onset was not available precluding use as inclusion criterion for control group patients. Although our integrated health system provides care for more than two-thirds of all COVID-19 hospitalizations in our region, it is possible that some patients may have been admitted to an ED or hospital outside of our system after testing, resulting in outcome misclassification.

Despite intentional and programmatic efforts at both the state and integrated healthcare network level to address healthcare disparities in MAb delivery, we did observe a significantly lower rate of MAb infusion to patients from communities of color. This can be partially understood by the dramatic intercurrent decline in the proportion of non-white patients among positive cases in Utah during the study period, from 56.3% in July 2020 to 27.5% by February 2021. However, this does not fully account for differences in the post-implementation group and highlights the ongoing need to address equity in healthcare access to COVID-19 care. Finally, our sample size was not sufficiently powered to detect rare adverse events or to make conclusions regarding comparative effectiveness between agents.

## Conclusion

In a real-world implementation targeting high-risk ambulatory COVID-19 patients within seven days of symptom onset, anti-SARS-CoV-2 monoclonal neutralizing antibody infusion appears to be well-tolerated and effective at preventing subsequent ED visits or hospitalizations. These data may help guide patient selection to maximize effectiveness of MAb therapy, an important consideration given current resource constraints of large-scale MAb deployment as part of public health efforts to decrease hospitalizations. Further, these data might guide implementation of future sustainable delivery models when Mab treatment is no longer government subsidized. Additional data from clinical trials on the impact on mortality, time to symptom resolution, and long-term sequelae are still needed to fully understand the benefit of these agents. Further study in larger real-world cohorts are necessary to better characterize rare infusion-related adverse events and to evaluate the comparative effectiveness of different Mab products. Linking genomic sequencing surveillance and monoclonal treatment data will also be important in future studies to inform differential effectiveness against emerging variant strains.

## Supporting information

STROBE Checklist

ICJME

## Data Availability

In order to protect patient privacy and comply with institutional data use policy, data used in this study are unavailable to upload to public servers. As required by the Intermountain Healthcare Institutional Review Board, data sharing agreement requests to access deidentified versions of the datasets generated and/or analyzed during the current study may be addressed to the Intermountain Office of Research.

## Author Contributions

Study concept: BJW, JB, WB, TV, IDP, SMB

Study design: BJW, WB, SMB, AMB

Data collection: NG, BJW

Statistical analysis: AMB, BJW, JB, IDP, SMB

Interpretation of results: All authors

Manuscript preparation: All authors

Critical review of the manuscript: All authors

## References

1. An EUA for Bamlanivimab-A Monoclonal Antibody for COVID-19. Jama. Dec 11 2020;doi:10.1001/jama.2020.24415

2. An EUA for casirivimab and imdevimab for COVID-19. Med Lett Drugs Ther. Dec 28 2020;62(1614):201–202.

3. Chen P, Nirula A, Heller B, et al. SARS-CoV-2 Neutralizing Antibody LY-CoV555 in Outpatients with Covid-19. The New England journal of medicine. Jan 21 2021;384(3):229–237. doi:10.1056/NEJMoa2029849

4. Gottlieb RL, Nirula A, Chen P, et al. Effect of Bamlanivimab as Monotherapy or in Combination With Etesevimab on Viral Load in Patients With Mild to Moderate COVID-19: A Randomized Clinical Trial. Jama. Feb 16 2021;325(7):632–644. doi:10.1001/jama.2021.0202

5. Weinreich DM, Sivapalasingam S, Norton T, et al. REGN-COV2, a Neutralizing Antibody Cocktail, in Outpatients with Covid-19. The New England journal of medicine. Jan 21 2021;384(3):238–251. doi:10.1056/NEJMoa2035002

6. Charlson ME, Pompei P, Ales KL, MacKenzie CR. A new method of classifying prognostic comorbidity in longitudinal studies: development and validation. J Chronic Dis. 1987;40(5):373–83. doi:10.1016/0021-9681(87)90171-8

7. Elixhauser A, Steiner C, Harris DR, Coffey RM. Comorbidity measures for use with administrative data. Med Care. Jan 1998;36(1):8–27. doi:10.1097/00005650-199801000-00004

8. Utah Crisis Standards of Care Monoclonal Antibody Allocation Guidelines (2020).

9. Webb BJ, Levin NM, Grisel N, et al. Simple Scoring Tool to Estimate Risk of Hospitalization and Mortality in Ambulatory and Emergency Department Patients with COVID-19. MedRxiv. 2021;doi:https://doi.org/10.1101/2021.02.22.21252171

10. Hernan MA, Robins JM. Using Big Data to Emulate a Target Trial When a Randomized Trial Is Not Available. Am J Epidemiol. Apr 15 2016;183(8):758–64. doi:10.1093/aje/kwv254

11. Rubin DB. Estimating causal effects from large data sets using propensity scores. Ann Intern Med. Oct 15 1997;127(8 Pt 2):757–63. doi:10.7326/0003-4819-127-8_part_2-199710151-00064

12. Austin PC, Stuart EA. Moving towards best practice when using inverse probability of treatment weighting (IPTW) using the propensity score to estimate causal treatment effects in observational studies. Stat Med. Dec 10 2015;34(28):3661–79. doi:10.1002/sim.6607

13. Chatellier G, Zapletal E, Lemaitre D, Menard J, Degoulet P. The number needed to treat: a clinically useful nomogram in its proper context. Bmj. Feb 17 1996;312(7028):426–9. doi:10.1136/bmj.312.7028.426

14. Wagner AK, Soumerai SB, Zhang F, Ross-Degnan D. Segmented regression analysis of interrupted time series studies in medication use research. J Clin Pharm Ther. Aug 2002;27(4):299–309. doi:10.1046/j.1365-2710.2002.00430.x

15. Libster R, Perez Marc G, Wappner D, et al. Early High-Titer Plasma Therapy to Prevent Severe Covid-19 in Older Adults. The New England journal of medicine. Feb 18 2021;384(7):610–618. doi:10.1056/NEJMoa2033700

16. Cevik M, Tate M, Lloyd O, Maraolo AE, Schafers J, Ho A. SARS-CoV-2, SARS-CoV, and MERS-CoV viral load dynamics, duration of viral shedding, and infectiousness: a systematic review and meta-analysis. Lancet Microbe. Jan 2021;2(1):e13–e22. doi:10.1016/S2666-5247(20)30172-5

17. Wang P, Liu L, Iketani S, et al. Increased Resistance of SARS-CoV-2 Variants B.1.351 and B.1.1.7 to Antibody Neutralization. bioRxiv : the preprint server for biology. Jan 26 2021;doi:10.1101/2021.01.25.428137

18. Starr TN, Greaney AJ, Dingens AS, Bloom JD. Complete map of SARS-CoV-2 RBD mutations that escape the monoclonal antibody LY-CoV555 and its cocktail with LY-CoV016. bioRxiv : the preprint server for biology. Feb 22 2021;doi:10.1101/2021.02.17.431683

19. Health UDo. Utah COVID-19 Variant Surveillance. Accessed March 15, 2021, https://coronavirus.utah.gov/case-counts/

