## Supplementary Material for "Real-World Effectiveness and Tolerability of Monoclonal Antibodies for Ambulatory Patients with Early COVID-19"

### eTable 1. Propensity Score Model

| Variable | Odds Ratio (95% CI) | P-value |
|---|---|---|
| Intercept | 0.08 (0.02, 0.32) | <.001 |
| **Age** | 1.01 (1.00, 1.02) | 0.005 |
| **Female** | 0.72 (0.60, 0.87) | <.001 |
| **Race** | | |
| Asian | 1.15 (0.27, 5.87) | 0.857 |
| Black or African American | 1.57 (0.32, 8.45) | 0.574 |
| Native Hawaiian or Pacific Islander | 1.81 (0.56, 8.11) | 0.371 |
| White | 1.80 (0.65, 7.46) | 0.329 |
| American Indian or Alaska Native | Ref | |
| **Hispanic or Latinx Ethnicity** | 1.01 (0.75, 1.34) | 0.972 |
| **Comorbidities** | | |
| Immunosuppression | 1.76 (0.82, 3.50) | 0.124 |
| Diabetes mellitus | 2.00 (1.67, 2.42) | <.001 |
| Coronary artery disease | 1.04 (0.78, 1.36) | 0.798 |
| Active Malignancy | 0.68 (0.30, 1.63) | 0.377 |
| Chronic pulmonary disease | 1.31 (1.09, 1.58) | 0.004 |
| Chronic kidney disease | 1.49 (1.20, 1.83) | <.001 |
| Chronic liver disease | 1.09 (0.89, 1.32) | 0.412 |
| Cerebrovascular disease | 1.11 (0.87, 1.40) | 0.412 |
| Hypertension | 1.84 (1.37, 2.50) | <.001 |
| Chronic neurological disease | 0.85 (0.65, 1.09) | 0.204 |
| Congestive heart failure | 1.36 (1.07, 1.72) | 0.011 |
| Cardiac arrhythmia | 1.16 (0.96, 1.40) | 0.132 |
| Obesity | 1.32 (1.08, 1.61) | 0.007 |
| **Testing location: Emergency Department vs Drive-up** | 0.42 (0.27, 0.63) | <.001 |
| **Symptoms at testing** | | |
| Fever | 0.97 (0.79, 1.19) | 0.781 |
| Cough | 1.06 (0.86, 1.32) | 0.563 |
| Short breath | 0.97 (0.78, 1.20) | 0.754 |
| Muscle or body aches | 1.27 (1.03, 1.57) | 0.025 |
| Runny nose | 0.95 (0.77, 1.16) | 0.611 |
| Loss of taste or smell | 0.79 (0.60, 1.03) | 0.089 |
| Sore throat | 1.15 (0.93, 1.41) | 0.185 |
| Diarrhea | 0.86 (0.66, 1.10) | 0.230 |
| **Patient Geographic Region of Residence** | | |
| Central rural regions | 0.04 (0.02, 0.09) | <.001 |

| | | |
|---|---|---|
| Davis county | 0.14 (0.07, 0.29) | <.001 |
| Eastern Salt Lake county | 0.14 (0.07, 0.29) | <.001 |
| Cache county | 0.08 (0.04, 0.16) | <.001 |
| Northern Utah county | 0.10 (0.05, 0.20) | <.001 |
| Southeastern Salt Lake county | 0.11 (0.05, 0.22) | <.001 |
| Southern rural regions | 0.03 (0.01, 0.08) | <.001 |
| Washington county | 0.07 (0.04, 0.15) | <.001 |
| Southern Utah county | 0.11 (0.05, 0.23) | <.001 |
| Southwestern Salt Lake county | 0.12 (0.06, 0.25) | <.001 |
| Wasatch Back | 0.10 (0.04, 0.21) | <.001 |
| Weber county | 0.07 (0.04, 0.15) | <.001 |
| Western Salt Lake county | 0.09 (0.04, 0.18) | <.001 |
| Cassia county (contrast) | 1 | |

**eFigure 1. Balance of clinical characteristics in the treated and contemporaneous control groups after inverse-probability of treatment weighting, by standardized mean differences**

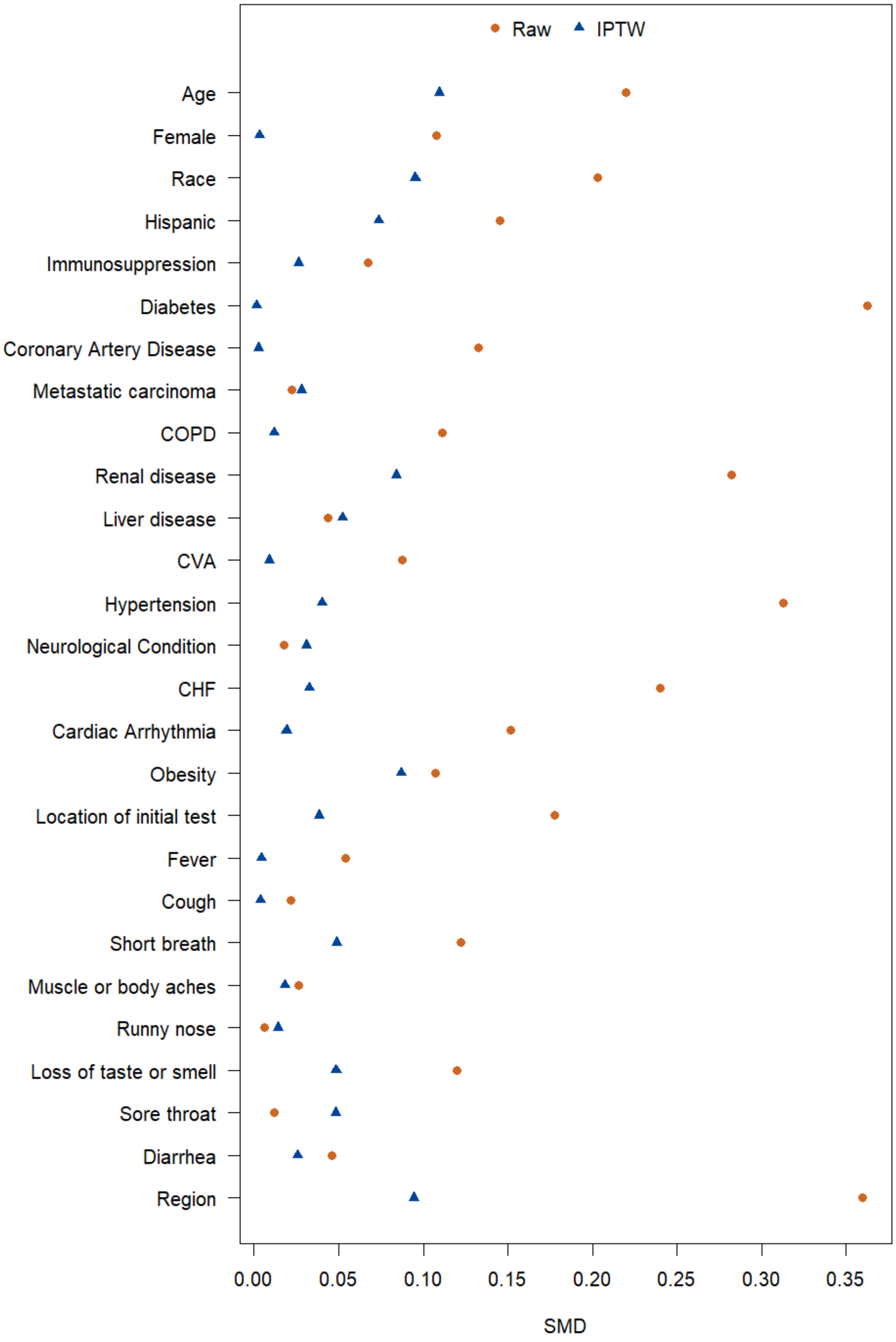

### eTable 2. Clinical features of patients treated at infusion centers only

| Variable | | Bamlanivimab | Casirivimab/ **Imdevimab** |
|---|---|---|---|
| N=272 | | 157 (57.7) | 115 (42.3) |
| Age, years, mean (SD) | | 64 (14) | 66 (15) |
| Female | | 38% (59) | 47% (54) |
| Race | | | |
| | American Indian or Alaska Native | 1% (1) | 0% (0) |
| | Asian | 1% (2) | 1% (1) |
| | Black or African American | 0% (0) | 1% (1) |
| | Native Hawaiian or Pacific Islander | 3% (5) | 1% (1) |
| | White | 91% (143) | 94% (108) |
| Hispanic or Latinx Ethnicity | | 12% (19) | 13% (15) |
| Communities of Color | | 18% (28) | 17% (19) |
| Total Comorbidities, median (IQI) | | 5 (3-6) | 4 (3-5) |
| | Immunosuppression | 6% (10) | 5% (6) |
| | Diabetes mellitus | 70% (110) | 63% (73) |
| | Coronary Artery Disease | 13% (21) | 14% (16) |
| | Active Malignancy | 3% (4) | 5% (6) |
| | Chronic pulmonary disease | 57% (90) | 57% (65) |
| | Chronic kidney disease | 32% (51) | 30% (35) |
| | Chronic liver disease | 28% (44) | 30% (34) |
| | Cerebrovascular disease | 21% (33) | 17% (20) |
| | Hypertension | 89% (140) | 89% (102) |
| | Chronic neurological disease | 13% (20) | 18% (21) |
| | Congestive heart failure | 24% (38) | 17% (19) |
| | Cardiac Arrhythmia | 46% (72) | 50% (57) |
| | Obesity | 68% (107) | 54% (62) |
| Outcomes | | | |
| | Emergency department visit (14 days) | 19 (12.1) | 9 (7.8) |
| | Hospital admission (14 days) | 7 (4.5) | 1 (0.9) |
| | Mortality (14 days) | 0 (0) | 0 (0) |
| | Composite outcome (14 days) | 19 (12.1) | 10 (8.7) |

**eTable 3. Segmented regression model with inverse-probability of treatment weighting estimating daily probability of the primary outcome.**

| Variable | Log Odds (95% CI) | P-value |
|---|---|---|
| Intercept | -1.266 (-1.423, -1.112) | <.001 |
| Baseline trend | -0.001 (-0.002, 0.001) | 0.428 |
| Level change | -0.075 (-0.237, 0.085) | 0.358 |
| Trend change | -0.007 (-0.010, -0.003) | <.001 |

**eTable 4. Multivariable logistic regression model comparing casirivimab/imdevimab versus bamlanivimab on the primary outcome.**

| Variable | Odds Ratio (95% CI) | P-value |
|---|---|---|
| Intercept | 0.00 (0.00, 10087114.62) | 0.411 |
| Female | 1.02 (0.45, 2.23) | 0.966 |
| Obesity | 0.97 (0.44, 2.23) | 0.936 |
| Secular trend in contemporaneous controls | 1.67 (0.37, 7.34) | 0.496 |
| Casirivimab/Imdevimab vs Bamlanivimab | 0.52 (0.17, 1.63) | 0.255 |
